# Reluctant Owners and Unwanted Guns: Exploring Motivations for Relinquishing Firearms at Gun Buybacks in Michigan

**DOI:** 10.64898/2026.03.29.26349650

**Authors:** David K. Humphreys, Deanna Giraldi, Sara Solomon, Elissa Trumbull, Douglas J. Wiebe

**Author notes:** **Corresponding author:** Professor David K. Humphreys, Department of Social Policy and Intervention, University of Oxford, 32 Wellington Square, Oxford, OX1 2ER.

## Abstract

**Background:** Firearms are frequently transferred through inheritance and other non-purchase pathways, leaving many individuals in possession of unwanted guns and limited options for safe disposal. This study examined the characteristics and motivations of individuals relinquishing firearms at community gun buyback and destruction events in Michigan to inform understanding of firearm divestment and disposal pathways.

**Methods:** We conducted an explanatory sequential mixed-methods study of six faith-based gun buyback and destruction events held in southeastern Michigan between June and October 2024. Quantitative surveys (n = 109) captured participant demographics and firearm characteristics. Follow-up qualitative interviews (n = 7) explored participants’ experiences and motivations using inductive–deductive thematic analysis.

**Results:** Across six events, 151 individuals relinquished 318 firearms, most of which were handguns. Nearly one third of participants disposed of firearms on behalf of others, and two thirds of personally owned guns had been obtained through non-purchase transfers, most commonly inheritance. Participants frequently expressed anxiety about storing unwanted firearms and relief after safe disposal. The most common motivations were concern about misuse (59%) and fear of theft (54%). Interviews identified five intersecting themes: inheritance and unwanted firearms, safety and family protection, evolving views on ownership, barriers to legal disposal, and emotional relief and closure after relinquishment.

**Conclusions:** Many individuals become firearm owners through inheritance or other non-purchase transfers rather than intentional acquisition. Their experiences reveal that unwanted firearms can generate sustained unease and moral responsibility, motivating voluntary divestment when safe, non-punitive options are available.

## INTRODUCTION

Findings from the 2018 Small Arms Survey estimated that over 393 million firearms are currently in circulation in the United States, representing the largest civilian firearm stock globally.^1^ At its present trajectory the U.S. firearm stock is estimated to reach 564 million firearms by 2034.^2^ Firearms are highly durable products, that can remain functional for decades, often outliving their original owners. As a result, efforts to limit firearm access to responsible and trained users may be undermined when weapons circulate through gifts or inheritance after owners die or become too frail to keep them. The accumulation and persistence of firearms in private homes has important implications for public safety across generations.

Firearms are frequently transferred between owners through pathways that are poorly understood. Estimates from the National Firearm Survey (NFS) suggest that a substantial proportion of owners acquire firearms through “non-purchase transfers” (i.e. gifts, inheritance, etc.).^3^ These findings suggest that a significant proportion of firearm transfers may be passive (i.e. via gifts, inheritance) rather than intentional (e.g. retail or gun-show purchases). This aligns with evidence showing that around five percent of firearm owners have attempted to relinquish a firearm within the past five years.^3,4^ Understanding how individuals come to possess and then divest of firearms is critical for designing policies that address the life cycle of civilian gun ownership. Without such insight, prevention efforts risk overlooking the social and structural pathways through which firearms circulate, accumulate, and persist in homes where they are neither wanted nor safely managed.^4,5^

In this study, we examine the perspectives of firearm owners seeking to relinquish firearms through community buyback and destruction events in Michigan. Although debates about the effectiveness of buyback programmes as a violence-prevention tool remain active,^6^ research has rarely examined the motivations or experiences of individuals who voluntarily relinquish firearms. We partnered with community organisations in southeastern Michigan in 2024 to document buyback implementation and to understand the circumstances and motivations of such individuals.

## METHODS

### Design

We used an explanatory sequential mixed-method design, with an initial quantitative phase examining the characteristics and motivations of buyback participants, followed by a qualitative phase to contextualise and elaborate on quantitative findings.^7^ Survey results informed the development of a topic guide for qualitative follow-up interviews.

### Programme implementation and setting

Between June and October 2024, six gun buyback and destruction events were organised in southeastern Michigan in partnership with local faith organisations. Events were community funded (for further details, see Appendix A) and operated by volunteers, with researchers from the University of Michigan and the University of Oxford present to observe and administer surveys.

### Patient and Public Involvement

Patients and members of the public were not involved in the design, conduct, reporting, or dissemination plans of this research.

### Data sources

We used three sources of data: (1) administrative data documenting basic donor and firearm characteristics compiled by the event organisers; (2) survey data collected from participating individuals during the events; and (3) qualitative data from seven in-depth interviews conducted with a sub-sample of survey participants.

Organisers routinely collected information from donors upon arrival, including date, donor demographics (e.g. age, address, etc.), firearm make and model, serial number, calibre, and compensation received ($50 for a long gun, $100 for a pistol, $200 for an assault rifle). These data served two purposes: (1) to document for internal reporting, and (2) to provide firearm-related information (e.g. serial numbers, calibre) to relevant authorities (e.g. law enforcement and the Bureau of Alcohol, Tobacco, Firearms and Explosives). These data were de-identified prior to transfer to the study team. The survey was hosted in Qualtrics survey software and completed on-site either via research team tablets or participants’ own devices using a QR code.^8^ It comprised four sections: (a) disposal information (firearm type, origin, reasons for relinquishment); (b) buyback information (how they learned about the event, prior attempts to relinquish a firearm); (c) social and demographic context (respondent demographics, community characteristics); and (d) follow up (willingness to be recontacted). The survey was designed to take less than 15 minutes to complete, aligning with the typical wait time for firearm destruction at events. We conducted seven follow-up semi-structured interviews with survey participants to further explore the motivations and context for relinquishing firearms (see Supplementary Appendix B for topic guide).

### Analysis

Descriptive statistics summarised donor demographics, firearm characteristics, and reported motivations. To contextualise our sample, we compared its demographic composition with the nationally representative National Firearm Attitudes and Behaviours Study (NFABs) survey,^9,10^ using Welch’s two-sample t-tests (for continuous variables) and chi-square tests (for categorical variables). All statistical analyses were conducted in R (version 4.3.2; R Core Team, 2023).

Interview transcripts were analysed thematically using a hybrid inductive–deductive approach.^11^ Initial codes were informed by survey findings (deductive), with additional codes generated inductively from the data. Two team members (DG, DH) coded transcripts independently, and themes were refined through team discussion. Qualitative analysis was conducted in NVivo.^12^ Results are presented in an explanatory sequence: quantitative findings appear first, followed by qualitative themes (with illustrative quotations) that contextualise and qualify the observed patterns. This explanatory sequential approach links quantitative patterns to the lived experiences of donors.

### Ethics

The study was approved by the University of Michigan, Medical School Institutional Review Board (HUM00262360, HUM00256770, HUM00262359).

## RESULTS

We present descriptive survey findings on donor characteristics, firearm types, and reported motivations, followed by the results from qualitative interviews to contextualise these patterns.

### Response rate and travel distance

Between July and November 2024, 151 individuals (hereafter ‘donors’) presented firearms for relinquishment and destruction at one of six events organised by faith groups in southeastern Michigan. Donors travelled a median of 15.6 miles (IQR 14.7; range 0–126.1) from their residences to the events. Of these 151 donors, 125 (82.7%) completed surveys and 109 (72.2%) provided full responses for analysis. Seven participants (6%) completed in-depth follow-up interviews.

### Demographics

Participants were diverse in sex, race and income (Table 1). Just over half (53.2%) were male, with an average age of 56.8 years. Compared with the national population of firearm owners, participants were more demographically diverse and less likely to reside in rural areas or be married (Table 1).

**Table 1:**
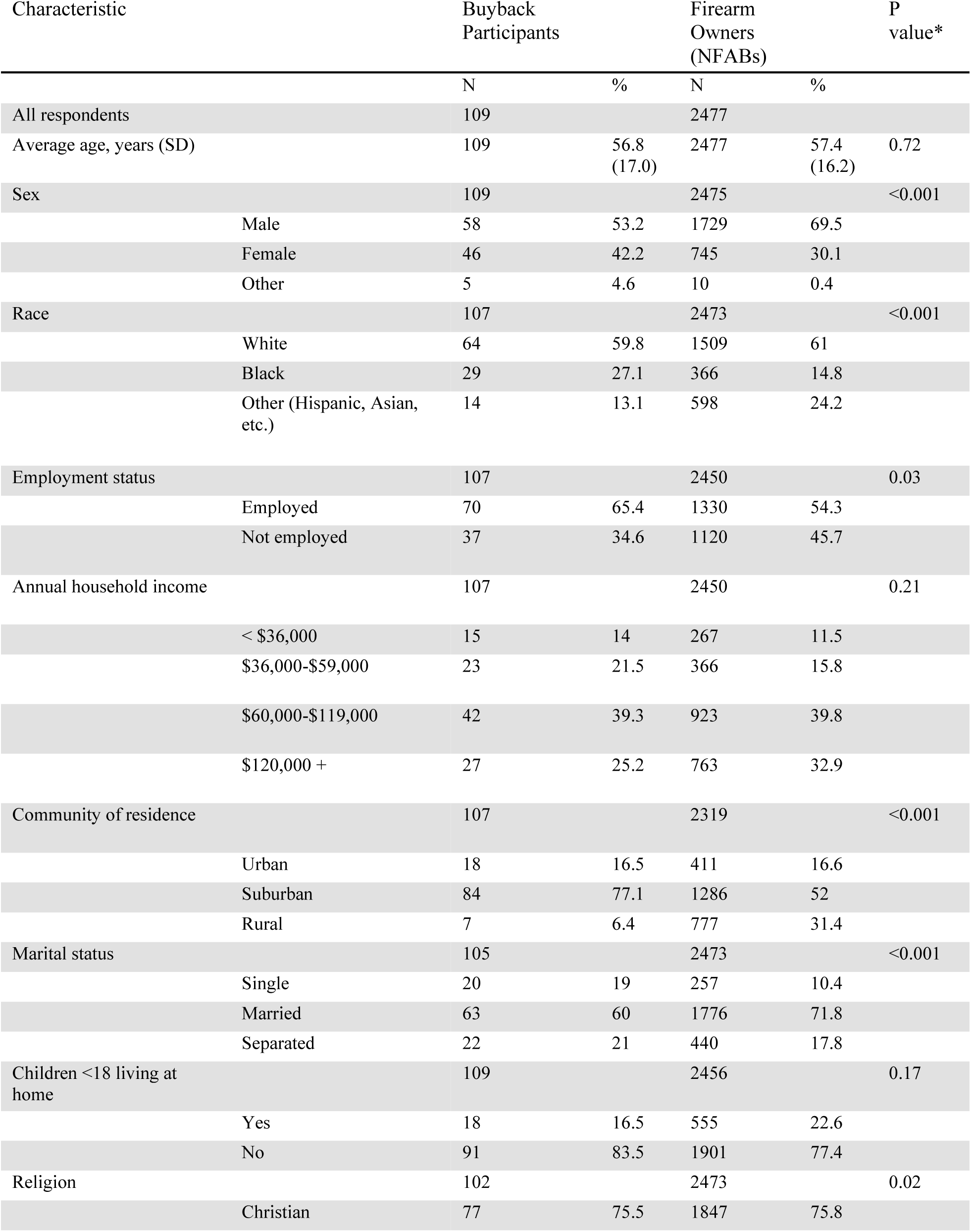
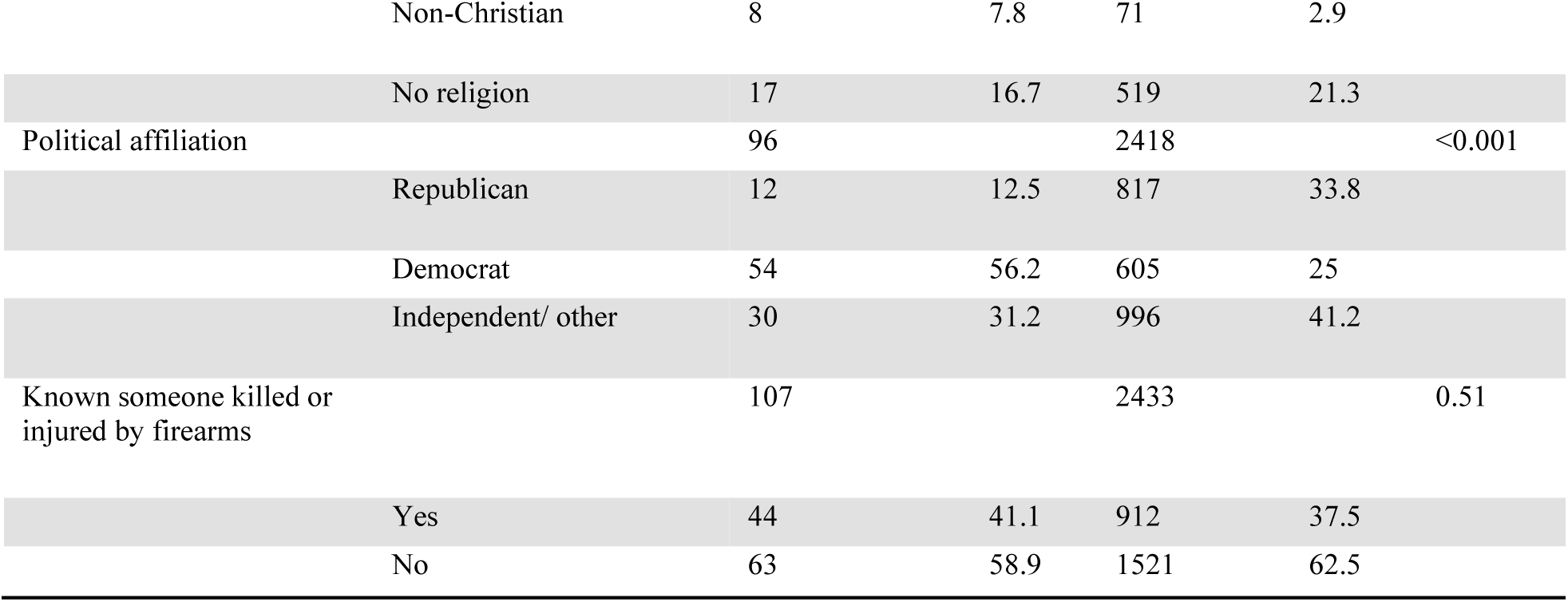
Characteristics of Survey Participants by Demographic Group Compared with Nationally Representative Firearm Owners.

**Table 2:** The Characteristics and Provenance of Relinquished Firearms*.

| Characteristic | N | % |
| --- | --- | --- |
| N relinquished by survey participants (% of total yield) | 174 | 54.7 |
| N firearms disposed per participant (n=109) |  |  |
| 1 firearm | 68 | 41.2 |
| 2-3 firearms | 36 | 21.2 |
| >3 firearms | 5 | 3 |
| Type of firearm donated by participants (n=165)* |  |  |
| Handgun | 104 | 63 |
| Long gun | 55 | 33.3 |
| Other | 6 | 3.6 |
| Missing | 9 | 5.5 |
| Who does the firearm belong to? (n=165) |  |  |
| Myself | 112 | 67.9 |
| Someone else | 53 | 32.1 |
| How did you come to own this firearm (n=112) |  |  |
| Purchased it | 36 | 32.1 |
| It was given to me | 76 | 67.9 |
| How was this firearm passed on to you? (n=76) |  |  |
| I inherited it | 51 | 67.1 |
| I traded it | 2 | 2.6 |
| It was a gift | 14 | 18.4 |
| Other | 9 | 11.8 |
| How long have you owned this firearm? (n=112) |  |  |
| < 1 year | 7 | 6.3 |
| 1-3 years | 29 | 25.9 |
| 3-5 years | 12 | 12 |
| > 5 years | 64 | 57.1 |
| What is the most important reason for owning this firearm? (n=112) |  |  |
| Protection | 33 | 29.5 |
| Target shooting | 21 | 18.7 |
| Hunting | 5 | 4.5 |
| Another reason | 13 | 11.6 |
| I never wanted this gun | 40 | 35.7 |

\*Participants were asked for detailed information for only the first three firearms they donated

### Firearms disposed

Drawing on administrative data from the six events, 151 individuals donated 318 firearms, all of which were destroyed on-site. Events yielded a mean of 53 firearms. Most firearms were handguns (59%), followed by long guns (35%, including seven fully-functional assault rifles), and other (unknown) types (6%, e.g. pellet or air guns). The median number of firearms donated per person was one, but 24.2% of donors relinquished more than one firearm. Remuneration was capped at $350 per donor and six donors declined compensation.

Survey participants reported donating 174 firearms–55% of the total yield. Nearly all survey participants (n=104, 95%) donated between one and three firearms, while five (5%) donated between four and six with data missing on nine. Consistent with the administrative records, most firearms donated by survey participants were handguns (63%) and a third (33%) were long guns.

Survey participants provided detailed information on up to three donated firearms. Most (67.9%) relinquished firearms they personally owned, while nearly one-third (32.1%) were donated on behalf of others, typically a parent (30%), partner (30%), or acquaintance (36%). Among firearms owned directly by donors, more than two-thirds (68%) had been acquired through non-purchase transfer (inheritance, gift, or trade). Inheritance accounted for 67% of the firearms acquired through non-purchase transfers, representing 29% of all firearms relinquished by survey participants.

Most relinquished firearms (57%) were acquired more than five years ago, with the majority of these obtained through non-purchase methods (Figure 1). Reasons for ownership further underscore the involuntary nature of many acquisitions: 29% were acquired for self-defense, 19% for sport, and 4% for hunting, and more than one-third (36%) firearms were never wanted.

**Figure 1:**
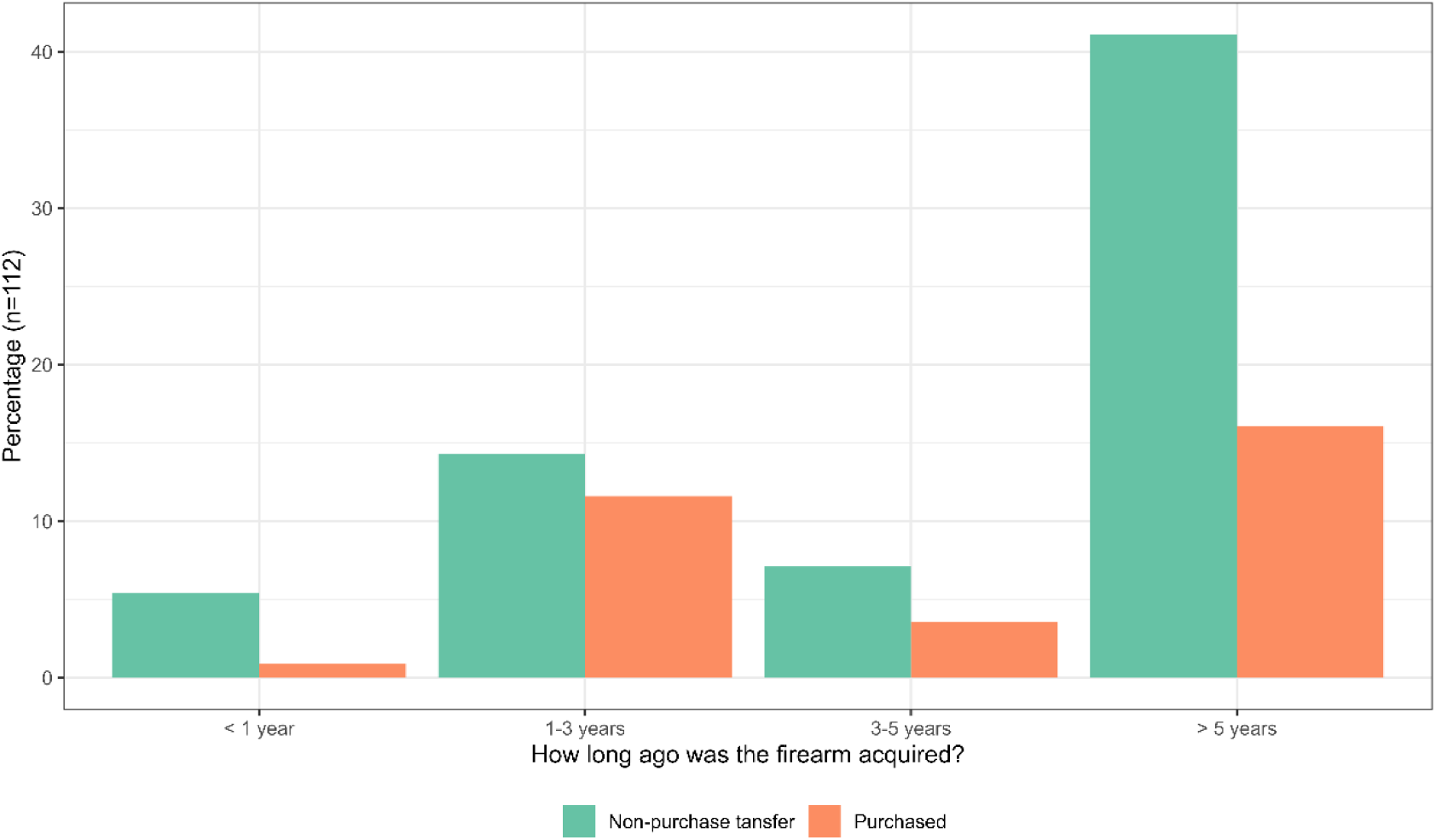
How long have donors owned these firearms by method of acquisition.

### Motivations

The most frequently endorsed reasons motivating participants to relinquish firearms were: (a) concern that others might misuse the firearm (59%) and (b) fear of theft (54%) (Figure 2). A majority disagreed with needing money from the buyback or concern for their own personal safety. About four in ten said the gift card was slightly important, while nearly half rated it unimportant. After the event, 43% reported they had no firearms and just over half retained between one and ten.

**Figure 2:**
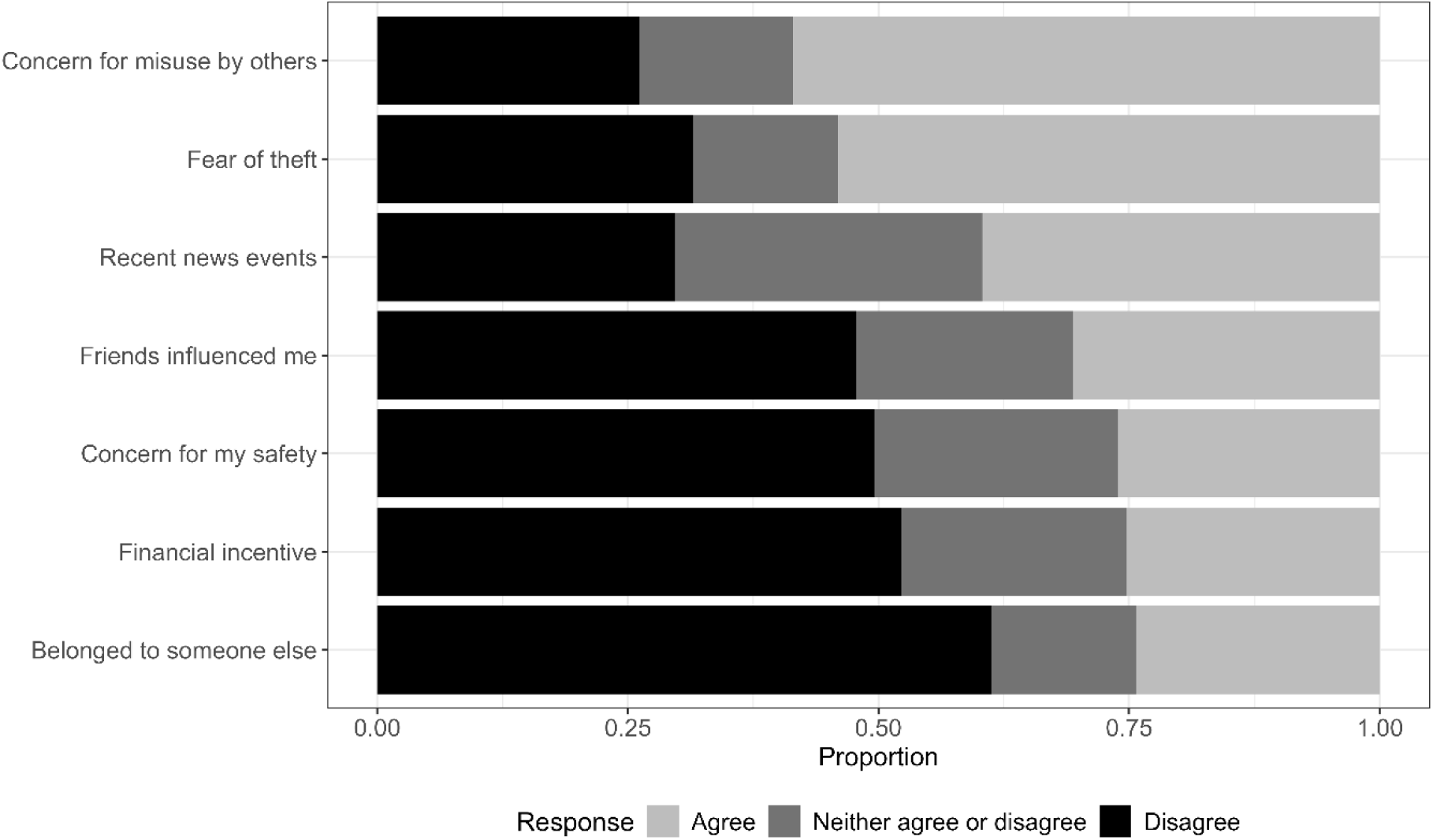
For what reasons are you disposing of a gun(s) today?

To contextualise these findings, we conducted in-depth interviews with seven survey participants. Thematic analysis identified five overarching themes: (1) inheritance and unwanted firearms, (2) safety, family, and protection, (3) evolving perspectives on gun ownership, (4) practical and legal barriers to disposal, and (5) emotional relief and closure. These themes highlight the social, cultural, and emotional dynamics shaping decisions to surrender firearms.

#### Inheritance and Unwanted Firearms

A dominant theme was the involuntary firearm acquisition, most often through inheritance. Participants (all names are pseudonyms) such as Alex, Malik, Michael, John, and Trisha described receiving weapons from deceased relatives, frequently characterised as unwanted or obsolete. Michael inherited firearms from two relatives; Trisha, serving as executor of her uncle’s estate, was tasked with liquidating his firearms despite her own anti-gun stance. Some acknowledged limited sentimental value but emphasised a desire to remove firearms from their households. Inherited firearms were framed less as heirlooms and more as burdensome objects conferring unwanted responsibility. This theme highlights the intergenerational circulation of firearms and the challenges of responsibly managing them in households where ownership is not otherwise normative (see Supplementary Appendix C for verbatim quotations).

> “So, two guns that I inherited and I didn’t really want…They just seemed a little, I don’t know, like dirty…It was like, oh, yeah, sure, I’ll take it, and then, after a while, I really don’t like this thing" (Michael)

#### Safety, Family, and Protection

Concerns about safety—particularly relating to children and mental health—were common motivations for divestment. Kevin linked his decision to create a gun-free household to the arrival of his first child, reframing firearms as preventable risks rather than protective assets. Trisha cited her children’s safety in the context of recurrent school shootings, while Christina described removing a firearm from a suicidal peer as part of a broader ethic of care. Participants described a tension between firearms as both symbols of protection and sources of vulnerability, often concluding that unintentional harm outweighed the protective benefits and highlighting the ongoing negotiation of risk, responsibility, and family wellbeing that owners manage. Alex, Malik, and Trisha further emphasised fears that unwanted firearms could be stolen, misused, or otherwise fall into the wrong hands—underscoring a strong sense of responsibility for preventing future harm.

> “Yeah. If you got an unwanted firearm or you don’t believe you’re gonna use it anymore, just get it out of circulation and give it to a gun buyback rather than putting it back on the streets…" (Malik)

#### Evolving Perspectives on Gun Ownership

Participants described shifts in their views on firearms shaped by life transitions and family backgrounds. For Kevin, parenthood prompted decisive change, as he moved from seeing guns as protective to recognising them as unnecessary risks. Trisha’s perspective was rooted in a childhood shaped by her father’s staunch opposition to guns, later reinforced by her own experiences of parenting amid recurrent local school shootings. Christina grew up in a household where firearms were normalised, recalling casual target practice as a child, yet she too ultimately came to question their protective value. Across these accounts, firearms were not treated as neutral possessions but as objects bound up with social, familial, and ethical meaning, continually re-evaluated as circumstances changed.

> “Once I had my son, my mind completely changed.” (Kevin)

#### Practical and Legal Barriers to Disposal

Participants encountered obstacles when seeking legal disposal options. Christina described being turned away by multiple police jurisdictions and left uncertain about the legality of possessing an unregistered weapon. Michael received contradictory instructions from different municipalities and worried that bringing an inherited, unregistered gun to the police could itself be a crime. John encountered resistance when attempting to register his father’s firearms, being told they could be confiscated. Alex found that neither local nor state police would accept his unwanted gun, leaving him at a loss until a buyback event was available. Both Kevin and Malik expressed discomfort with taking firearms to the police, citing fears of being misunderstood, treated with suspicion, even preferring to throw a gun away rather than carry it into a station. Buyback programmes were consistently described as simple, accessible, and non-punitive alternatives. These accounts underscore how gaps in firearm governance—conflicting guidance, bureaucratic obstacles, and limited lawful disposal pathways—leave owners discouraged from compliance even when seeking to act responsibly.

> "I tried to give it to the local police and State police, and they did take the bullets but they would not take the gun. So, I just didn’t know what to do with it.” (Alex)

#### Emotional Relief and Closure

Participants consistently described feelings of relief and resolution following firearm disposal. Kevin reported pride and peace of mind in removing risk from his home, while Trisha spoke of the immediate sense of safety and calm that came from clearing her house of guns she had long felt uneasy about. Malik emphasized relief in finally relinquishing his father’s handgun, with the knowledge that it was destroyed easing his conscience. Alex described closure after years of anxiety about storage and theft, noting the satisfaction of being “completely gun free.” Christina recounted a collective effort to remove a firearm from a suicidal peer, framing disposal as both a personal and communal responsibility. These narratives suggest that participation was not merely pragmatic but also a symbolic resolution of ethical, emotional, and familial tensions associated with firearm possession.

> "I feel so much safer without a gun. Even having those two guns…They were in my house, but they were in our attic where nobody could really access them. I still didn’t feel safe having them up there because we have two small children…I don’t know what I thought could happen, but you know just the thought for me. It was like, get these out of my house as soon as possible. I don’t want them. It makes me scared and nervous…It was a really good feeling, driving away and knowing like that kind of is gone, you know." (Trisha)

Taken together, these findings demonstrate that firearm relinquishment at community buyback events is shaped by a combination of structural, social, and personal factors. While survey data highlighted safety concerns and involuntary ownership as prominent motivations, interviews revealed the deeper familial, cultural, and emotional dynamics underlying these decisions.

## DISCUSSION

This study examined firearm owners who voluntarily relinquished unwanted guns at community buyback and destruction events in southeastern Michigan. Across six events, more than 150 individuals surrendered 318 firearms, mostly handguns. Two-thirds of donors agreed to participate in surveys and interviews about the firearms they were relinquishing and their motivations for doing so. Nearly one third of relinquished firearms were donated on behalf of others (e.g. parents, partners, acquaintances) and most personally owned firearms were acquired through non-purchase transfers, typically inheritance. Participants often retained these firearms for many years before being able to dispose of them. These patterns reveal how unwanted firearms persist in civilian circulation, highlighting an overlooked dimension of population-level exposure to firearm risk. Whether acquired intentionally or not, many participants expressed anxiety and discomfort about storing firearms they did not want and relief when safe disposal became possible.

Debates about gun buybacks often focus narrowly on their effectiveness in reducing crime rates.^6,13^ Yet this framing misses a broader public health function: providing one of the few accessible, non-punitive pathways for firearm divestment. Participants emphasized that while purchasing firearms in the U.S. is straightforward, pathways for safe and legal disposal remain sparse.^14,15^ For many, the presence of an unwanted firearm represented an enduring safety concern—posing risks of theft, suicide, or unintentional harm within families. Decisions to relinquish firearms reflected not only personal values but also a sense of collective responsibility for preventing harm.

Emerging evidence links voluntary firearm divestment with substantial reductions in suicide risk, underscoring divestment as an underused injury-prevention strategy.^5^ Our findings extend this evidence by identifying a distinct group of reluctant firearm owners—individuals who acquire guns through inheritance or other non-purchase transfers, or who later regret ownership. Their experiences complicate assumptions that gun ownership is always voluntary or desired and expose a policy gap in firearm governance: the absence of routine accessible disposal mechanisms. For these individuals, community buybacks offered an essential opportunity to reduce household and community risk in the absence of established disposal infrastructure. Delays many reported before successfully relinquishing a firearm highlight the need for permanent divestment options as part of comprehensive firearm-injury prevention.

This study offers rare insights into the perspectives of individuals relinquishing firearms, a population often overlooked in firearm research and policy debates.^4,16^ Although exploratory, geographically limited and based on a small cross-sectional convenience sample, our findings suggest directions for future research on the life course of civilian firearms: who they are passed to, how recipients are affected, and under what conditions owners seek to divest. Further work is needed to understand how reluctant ownership operates across different populations and what practical support might facilitate safe firearm disposal. Motivations for divestment may differ substantially in communities where the absence of a firearm is perceived as a vulnerability to predatory crime.^17^ The implications of this study may take a different shape in urban or high-risk contexts, underscoring the need for comparative research.

## CONCLUSION

Firearms routinely enter households through non-purchase pathways and may remain for many years despite generating anxiety and perceived risk. While buyback programmes alone may not directly prevent violence, they address a crucial public health need by offering safe and accessible disposal options to individuals seeking to divest. Expanding these models—through permanent community collection points, integration with healthcare screening, and partnerships with faith or civic organisations—could institutionalise safe disposal as a core harm-reduction strategy. Recognising firearm relinquishment as part of the public-health continuum of ownership, storage, and disposal will help prevent injuries and deaths associated with unwanted firearms. This is particularly urgent in the U.S. context, where the civilian stock of firearms continues to grow,^2^ and concentrate in older age groups,^18^ trends that are likely to increase the likelihood of unwanted inheritance.

## Data Availability

The data underlying this study are not publicly available due to the sensitive nature of the subject matter and the risk of participant identification. De-identified data may be made available from the corresponding author upon reasonable request, subject to institutional and ethical approvals.

## COMPETING INTERESTS

The authors declare no competing interests.

## ACKNOWLEDGEMENTS

We would like thank Jill Soloman, Taylor Hautala, Nichole Burnside for their contributions to data collection, event logistics and coordination. We are also grateful to members of the Institute for Firearm Injury Prevention, in particular, Jessica Roche, Sonia Kamat, Haley Crimmins, Dr. Rebeccah Sokol and Dr. Dan Lee who advised on survey development. We also acknowledge the support of Fr. Chris Yaw and partners at St. David’s Episcopal Church and collaborating congregations of the Trinity Gun Buyback and Destruction Collaboration for facilitating access to the study sites. Finally, we are grateful to the participants who shared their experiences and to the volunteers and staff whose efforts made this project possible.

## FUNDING

This work was supported by exploratory research grants from both the University of Michigan Firearm Injury Prevention (IFIP) and University of Michigan Injury Prevention Center (IPC).

## SUPPLEMENTARY APPENDIX A

Budget Breakdown of Buyback and Destruction Events (June-October 2024)

**Budget Breakdown of Buyback and Destruction Events (June-October 2024)**
| **Expenditure category** | **Amount**<br>**(USD)** | **% of Total**<br>**Budget** | **Description** |
| --- | --- | --- | --- |
| Gift cards for participants | \$22,267 | 32.9 | Remuneration for firearm donors |
| Insurance liability | \$21,584 | 31.9 | Event insurance coverage |
| Equipment | \$14,000 | 20.7 | Circular saws, blades, tables, generators |
| Advertising and promotion | \$4,600 | 6.8 | Flyers, local media, community outreach |
| Private security | \$3,800 | 5.6 | Hired security personnel |
| Art supplies | \$1,000 | 1.5 | For community awareness/ engagement art |
| Transportation | \$200 | 0.3 | Moving equipment and materials |
| Total | \$67,451 | 100 | |

## SUPPLEMENTARY APPENDIX B

This guide was used to structure in-depth interviews with participants who disposed of firearms at buyback events. It illustrates the range of topics explored; interviews were conducted flexibly, with follow-up questions depending on participants’ responses.

Topics covered:

1. **Background**

- Participant’s personal history and family background.
- Experiences growing up around firearms.
2. **Firearm(s) brought to the event**

- Origin and history of the firearm(s).
- How and why they came into the participant’s possession.
- Role (if any) of the firearm in their life.
3. **Other firearms in the household**

- Ownership of other guns by participant or household members.
- Feelings about living in a home with or without firearms.
4. **Decision to dispose**

- Motivations for relinquishing the firearm(s).
- Key influences (safety concerns, family, life events, personal values).
- Emotional and practical considerations.
5. **Process of disposal**

- Why the buyback event was chosen over other options.
- Experience of the disposal process.
- Role of law enforcement, services, or community organizations.
6. **Support and barriers**

- Information, advice, or assistance received.
- Gaps in support or challenges encountered.
- Suggestions for improving services.
7. **After-effects and reflections**

- Feelings after disposal.
- Impact on sense of safety, identity, and well-being.
- Changing views on gun ownership and gun policy.
8. **Final reflections**

- Additional thoughts or messages for policymakers or community member

## SUPPLEMENTARY APPENDIX C

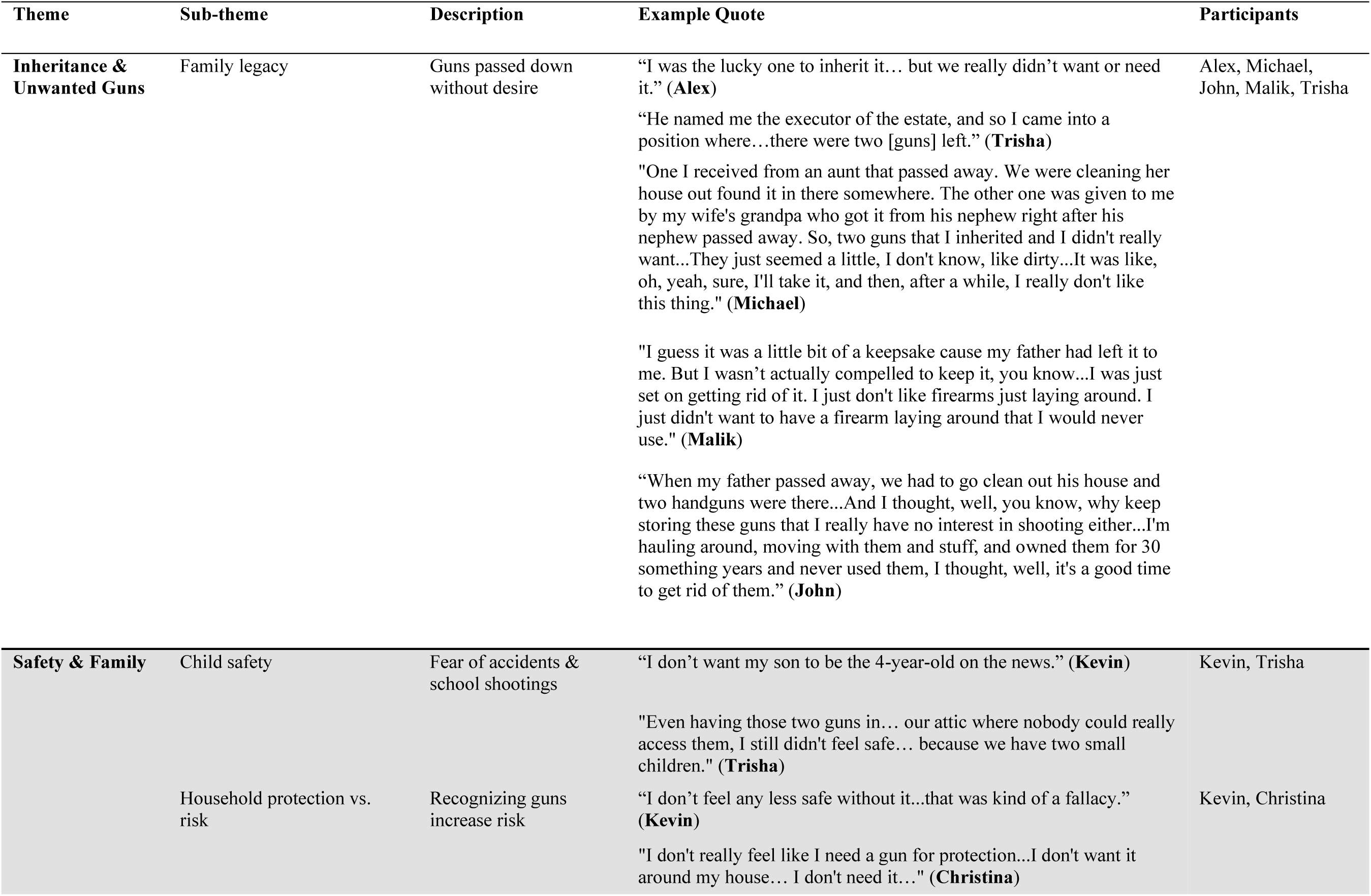
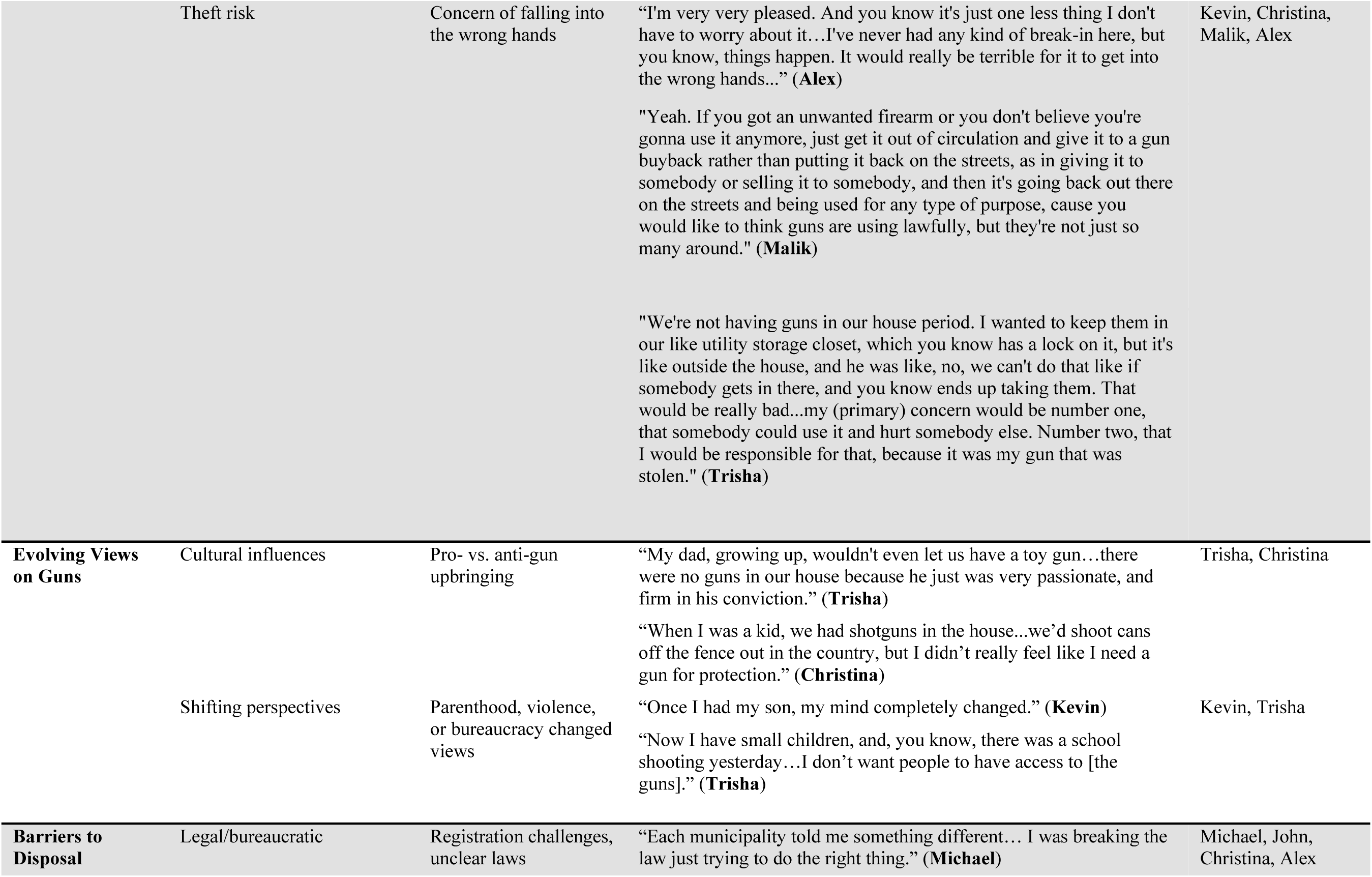
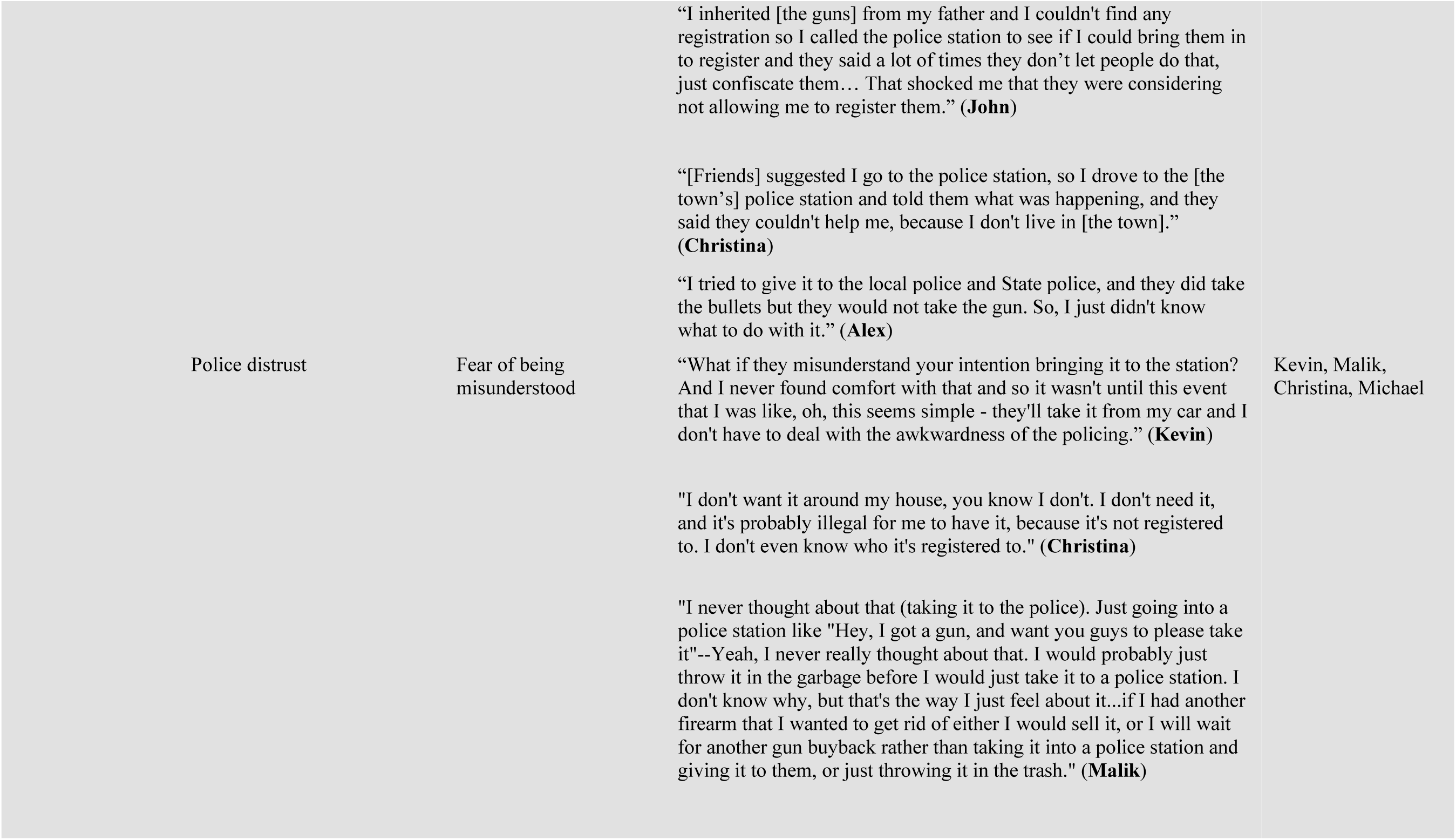
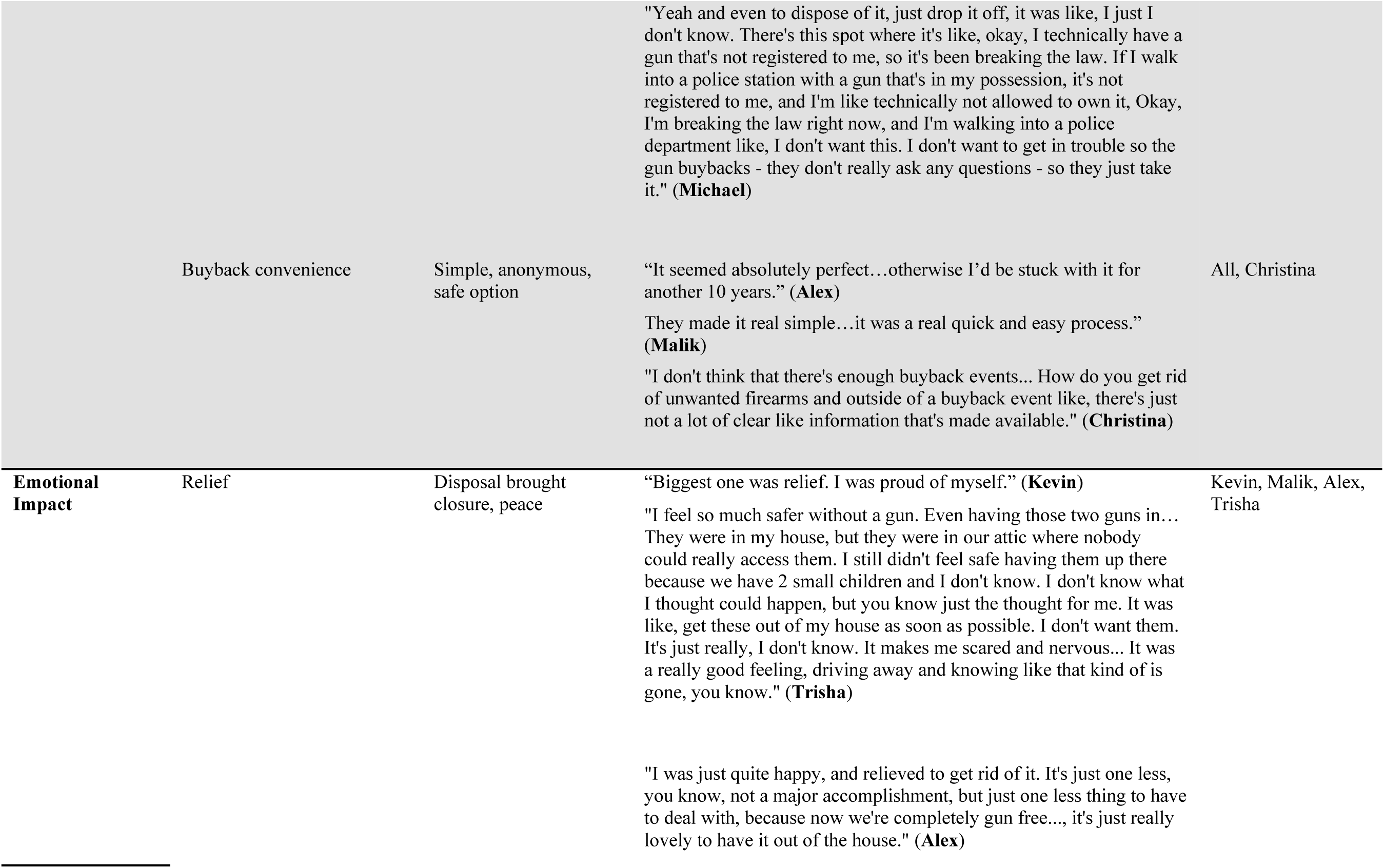
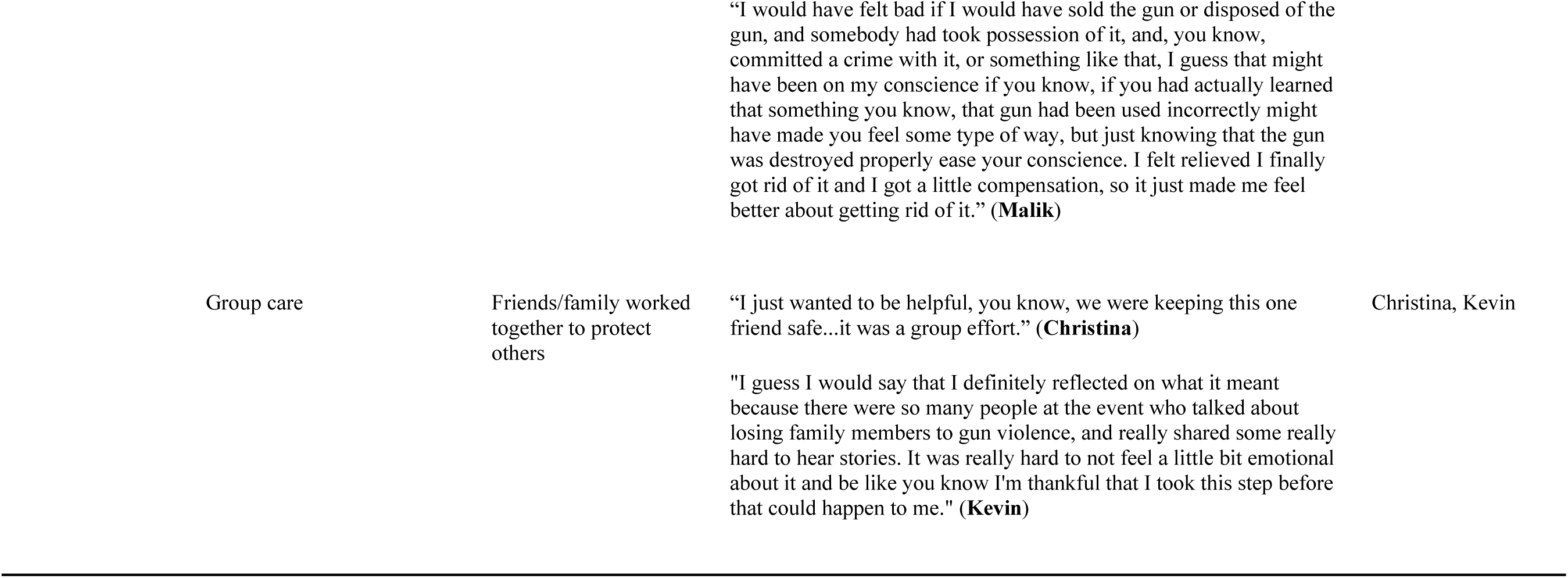

## Notes

### Competing Interest Statement

The authors have declared no competing interest.

### Author Declarations

Ethical approval for this study was granted by the University of Michigan Medical School Institutional Review Board (HUM00262360, HUM00256770, HUM00262359)

